# Development and Validation of a Diagnostic Nomogram to Predict COVID-19 Pneumonia

**DOI:** 10.1101/2020.04.03.20052068

**Authors:** Zhiyi Wang, Jie Weng, Zhongwang Li, Ruonan Hou, Lebin Zhou, Hua Ye, Ying Chen, Ting Yang, Daqing Chen, Liang Wang, Xiaodong Liu, Xian Shen, Shengwei Jin

## Abstract

**Background:** The COVID-19 virus is an emerging virus rapidly spread worldwide This study aimed to establish an effective diagnostic nomogram for suspected COVID-19 pneumonia patients.

**METHODS:** We used the LASSO aggression and multivariable logistic regression methods to explore the predictive factors associated with COVID-19 pneumonia, and established the diagnostic nomogram for COVID-19 pneumonia using multivariable regression. This diagnostic nomogram was assessed by the internal and external validation data set. Further, we plotted decision curves and clinical impact curve to evaluate the clinical usefulness of this diagnostic nomogram.

**RESULTS:** The predictive factors including the epidemiological history, wedge-shaped or fan-shaped lesion parallel to or near the pleura, bilateral lower lobes, ground glass opacities, crazy paving pattern and white blood cell (WBC) count were contained in the nomogram. In the primary cohort, the C-statistic for predicting the probability of the COVID-19 pneumonia was 0.967, even higher than the C-statistic (0.961) in initial viral nucleic acid nomogram which was established using the univariable regression. The C-statistic was 0.848 in external validation cohort. Good calibration curves were observed for the prediction probability in the internal validation and external validation cohort. The nomogram both performed well in terms of discrimination and calibration. Moreover, decision curve and clinical impact curve were also beneficial for COVID- 19 pneumonia patients.

**CONCLUSION:** Our nomogram can be used to predict COVID-19 pneumonia accurately and favourably.

## Introduction

Corona Virus Disease 2019 (COVID-19) pneumonia is confirmed to be infected with a novel coronavirus which is a β coronavirus that belongs to the family Coronaviridae. It has spread rapidly throughout Wuhan (Hubei province) to other provinces in China and around the world. ^1,2^ On January 30, 2020, the World Health Organization declared COVID-19 was a public health emergency of international concern (PHEIC). As of March 13, 2020, a total of 51767 laboratory-confirmed patients and 1775 deaths have been documented outside China. This pandemic has been disastrous for people all over the world, and many countries are failing to control its spread. The main reason was lack of rapid response which is depend on very early detection and diagnosis^3^.

As with all infectious diseases, the early and reliable diagnosis is key to block COVID-19 transmission. However, COVID-19 pneumonia has a wide range of clinical manifestations, such as fever, cough, fatigue, pharyngeal pain, etc^4^, Many patients have mild symptoms or asymptomatic in the early stage. Due to lack of specific clinical symptoms and signs, it is easy to be ignored.^5,6^ Real-time polymerase chain reaction (PCR) has become an important tool in the diagnosis of many infectious diseases^7,8^. However, it has several disadvantages, including long detection time, cumbersome steps, and high cost. Even the sensitivity of the COVID-19 nucleic acid detection is low. In addition, COVID-19 nucleic acid detection kits are still insufficient in many epidemic countries now. Detection of viral nucleic acid may not be the only ideal diagnostic method during the early stages of this epidemic outbreak.

Given the rapid spread of COVID-19 and low detection rate by pharyngeal swab COVID-19 nucleic acid test. It is also believed that throat swab samples are not suitable for the detection of SARS coronavirus RNA^9^. The development of a diagnostic method with decreased complexity and expense are urgently needed to facilitate timely intervention.^10^ Previous studies with significantly large sample sizes have been done to delineate the epidemiological and clinical characteristics of COVID- 19 pneumonia.^11^ Although some studies have shown that lung computed tomographic (CT) plays an important role in early diagnosis^12,13^. However, there is still a lack of systematic and standard imaging diagnostic criteria for COVID-19 pneumonia. To this end, 294 suspected COVID-19 pneumonia patients were included in our study. Trying to establish a diagnosis model to quickly identify COVID-19 pneumonia from suspected COVID-19 pneumonia.

## Methods

### Study design and participants

Two cohorts of adult suspected COVID-19 pneumonia patients were included in this retrospective study. The patients in primary cohort were from the Second Affiliated Hospital and Yuying Children’s Hospital of Wenzhou Medical University (Wenzhou, China), and the patients in validation cohort were from the People’s Hospital of Yueqing (Yueqing, China). We relied on new coronavirus pneumonia control and prevention plan (trial version 6) to identify suspected COVID-19 pneumonia patients. Suspected diagnostic criteria are as follows:1) epidemiological history. 2) fever and / or respiratory symptoms. 3) suspected COVID-19 pneumonia imaging features. 4) normal range or decrease of white blood cell (WBC) count or lymphocyte at beginning of disease. Patients who have epidemiological history + any two clinical symptoms or no epidemiological history + three clinical symptoms can be diagnosed as suspected COVID-19 pneumonia. All patients with suspected COVID-19 pneumonia were taken throat swab samples or sputum sample at admission (at least twice samples were taken, at least a 24 h apart) and stored in virus medium, which were transported to Wenzhou Center for Disease Control and Prevention (CDC) for COVID-19 diagnosis. The study was approved by the Ethics Committee of the Second Affiliated Hospital and Yuying Children’s Hospital of Wenzhou Medical University.

### Data collection

Demographic data, epidemiological history, comorbidity, vital signs, clinical symptoms, laboratory indicators including WBC count, neutrophils count, lymphocyte count, hemoglobin, platelet and C-reactive protein (CRP) and chest CT imaging features were collected. The data of laboratory indicators and chest CT imaging features were the first recorded data.

### Image acquisition and analysis

All chest CT images features in two cohorts were analyzed by four chest radiologists (two radiologists for one cohort) with least 10 years of experience in chest CT imaging and all decisions were reached by consensus. CT images features mainly include four parts: (1) lesion distribution, such as bilateral lower lobes, multiple lobes, periphery distribution (2) lesion patterns, such as patchy or large patchy distribution, wedge-shaped or fan-shaped lesion parallel to or near the pleura, crazy paving pattern, (3) lesion density, such as ground glass opacities, consolidation, cavitation, (4) other signs in the lesion, such as lung nodule, nodules halo sign, subpleural nodules, centrilobular nodules, other nodules, pleural effusion, air bronchogram sign, bronchiectasis, fibrotic proliferation, bronchial wall thickening, tree-in-bud pattern, white lung (Figure 1).

**Figure 1:**
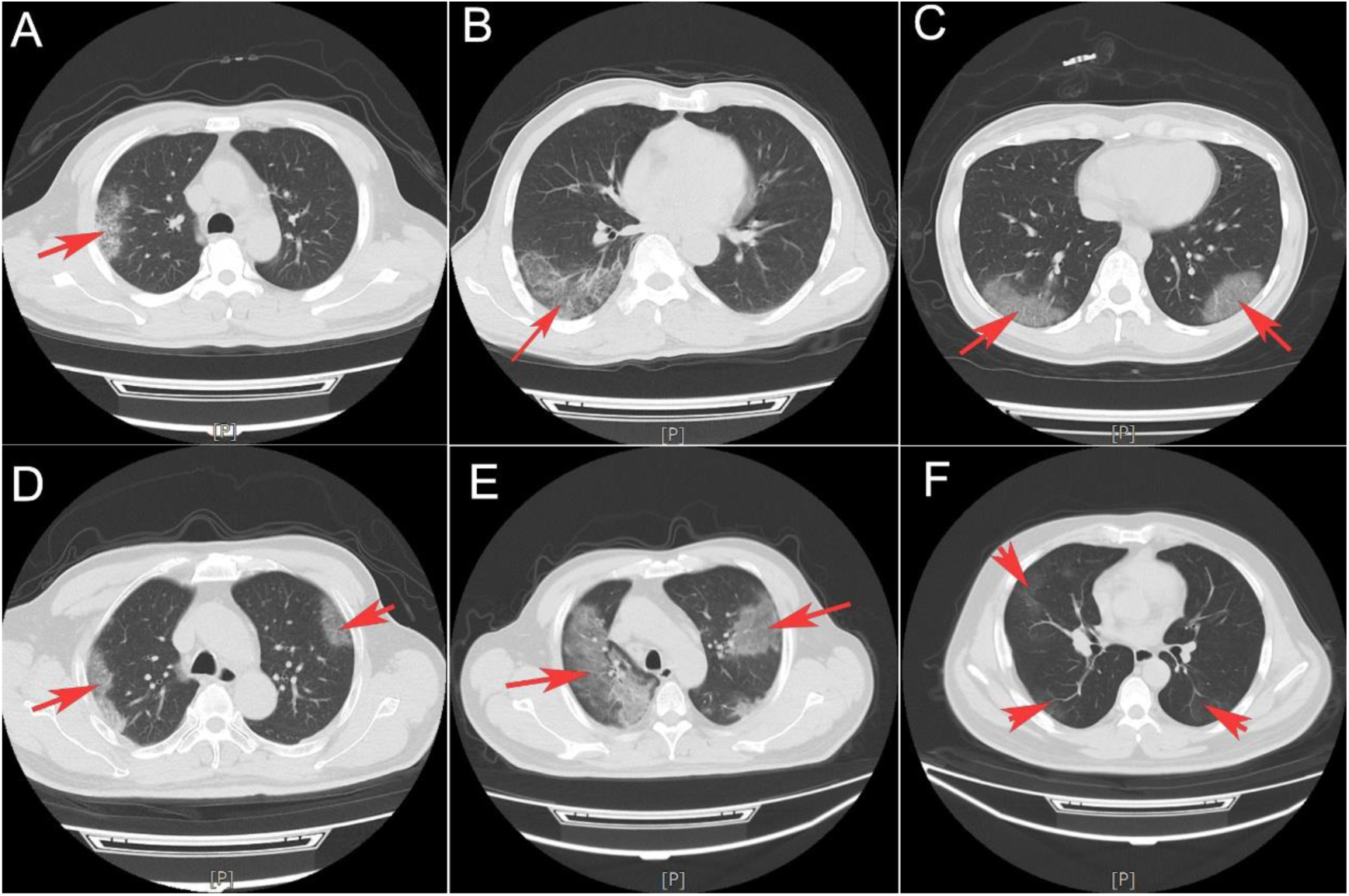
Classical COVID-19 pneumonia CT imaging features. Crazy paving pattern (A and B), wedge-shaped or fan-shaped lesion parallel to or near the pleura (C and D), Ground glass opacities (E and F).

### Statistical analysis

Data were presented as mean ± standard deviation (SD) and median (IQR) for continuous variables with normal or non-normal distribution and the frequency (proportion) for categorical variables. The Student t-test, Mann-Whitney U test, chi-squared test, or Fisher’s exact test was performed where appropriate.

For the development of the nomograms, the least absolute shrinkage and selection operator (LASSO) method was performed to identify potential significant predictors from the primary cohort. Predictive variables that were considered clinically relevant (based on our clinical experience and literature report), and that showed statistical relationship in LASSO method were entered into multivariate logistic regression model^14^. The Variance Inflation Factor (VIF) was used to identify variables collinearity before the model estimation. According to the Akaike information criterion (AIC) and clinically relevant variables, we established the final multivariable models. The ‘rms’ package was used for nomogram and calibration curve^15^. The accuracy of the nomogram to predict the COVID-19 pneumonia were quantified using C-statistic, the calibration of the model is assessed by the calibration curves in the primary cohort and validation cohort. Moreover, we performed decision curve analysis (DCA) and clinical impact curve by quantifying the net benefits and cost benefit ratio to assess the clinical value of the model^16^.

We did the statistical analyses and figures production using R software (version 3.6.1). All statistical tests were two-sided, differences of P < 0.05 were considered statistically significant.

## Results

The primary cohort consisted of 178 suspected COVID-19 pneumonia patients who received treatment in our hospital between Jan 25, 2020, and Mar 3, 2020. 89 patients were diagnosed with COVID-19 pneumonia (7 COVID-19 pneumonia patients were negative or weakly positive in initial viral nucleic acid detection). The validation cohort consisted of 116 suspected COVID-19 pneumonia patients. 68 patients were diagnosed with COVID-19 pneumonia (no COVID-19 pneumonia patients were negative or weakly positive in initial viral nucleic acid detection). No deaths occurred in the two cohorts. Patient characteristics, clinical symptoms, laboratory findings and chest CT imaging features were show in Table 1.

**Table 1:**
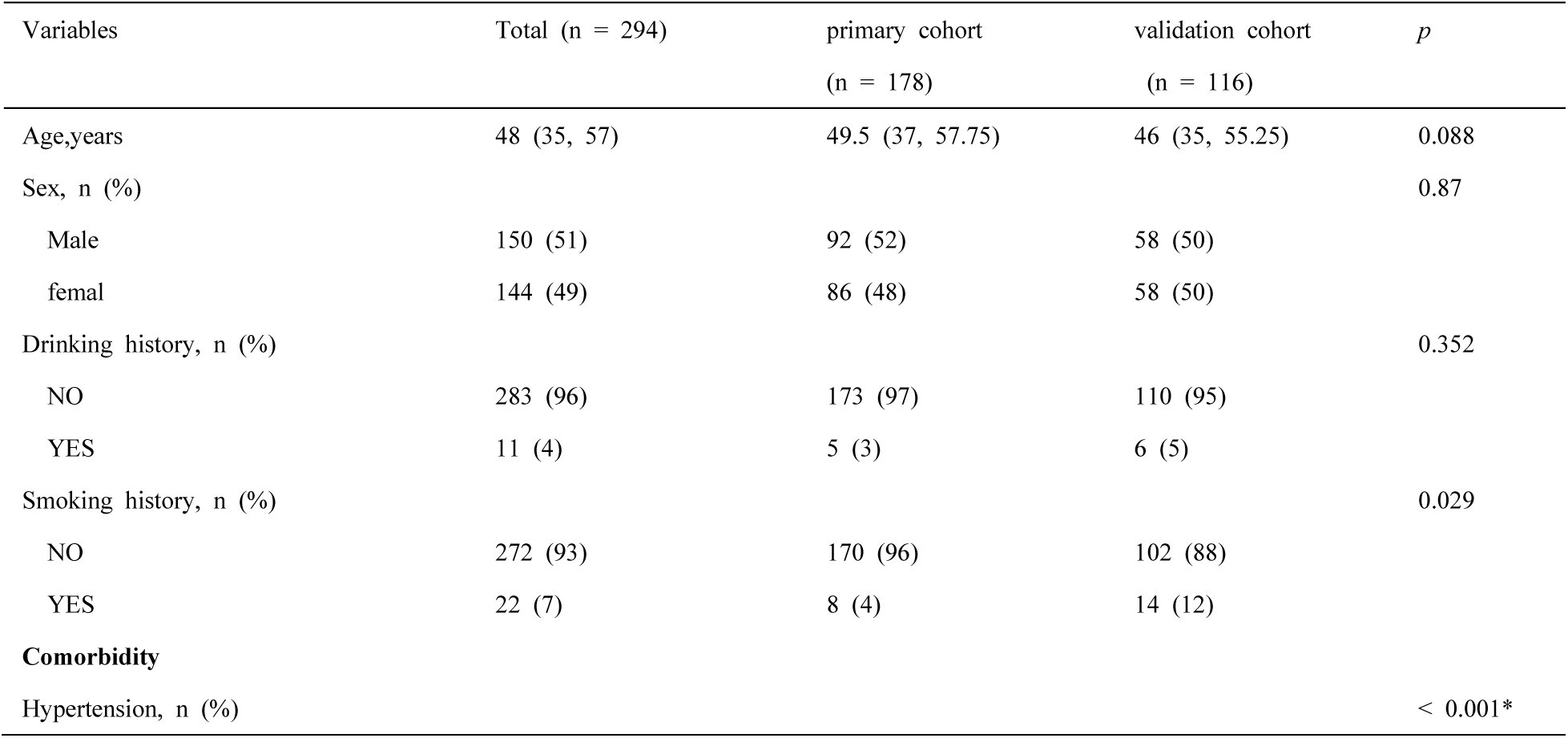
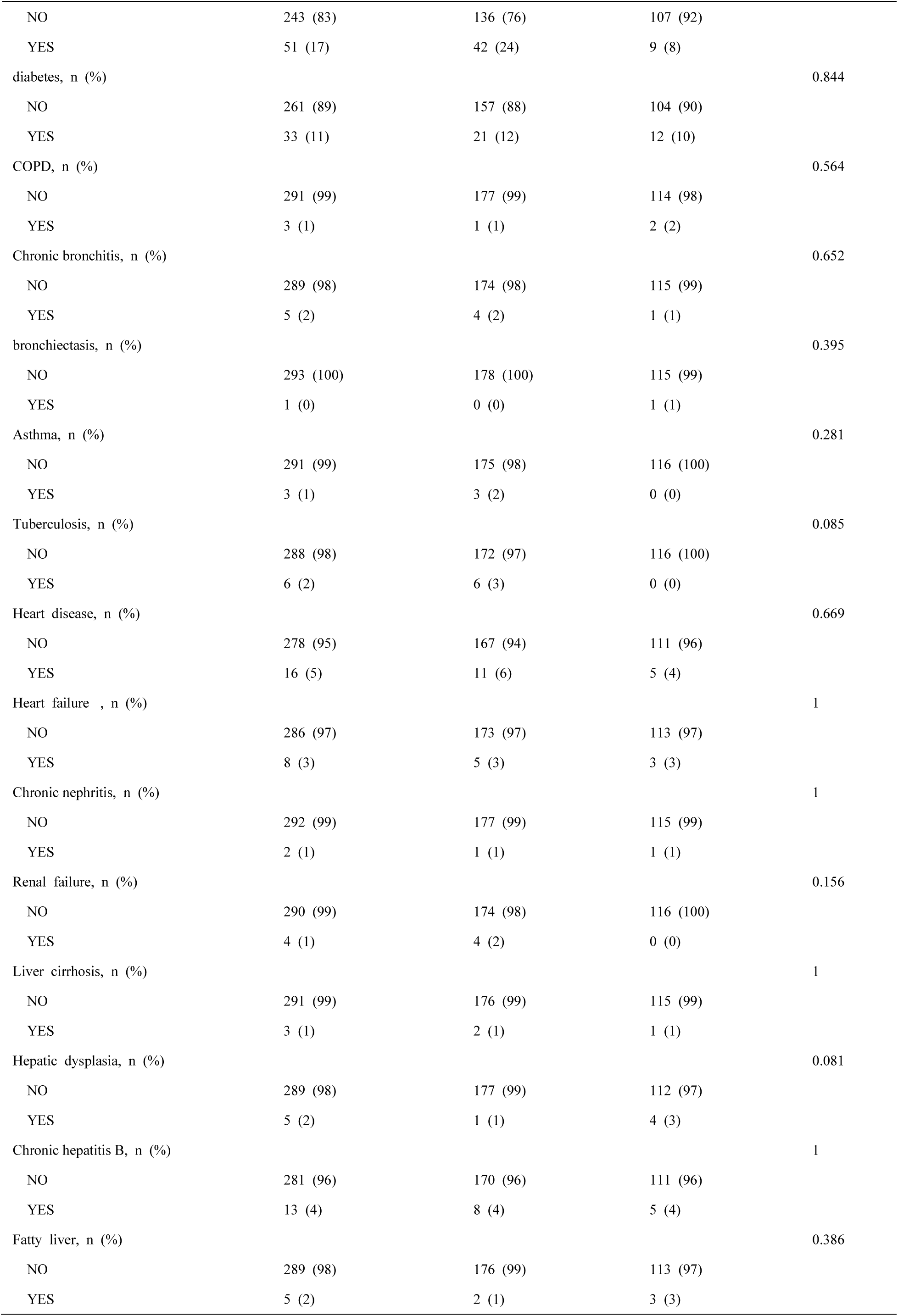
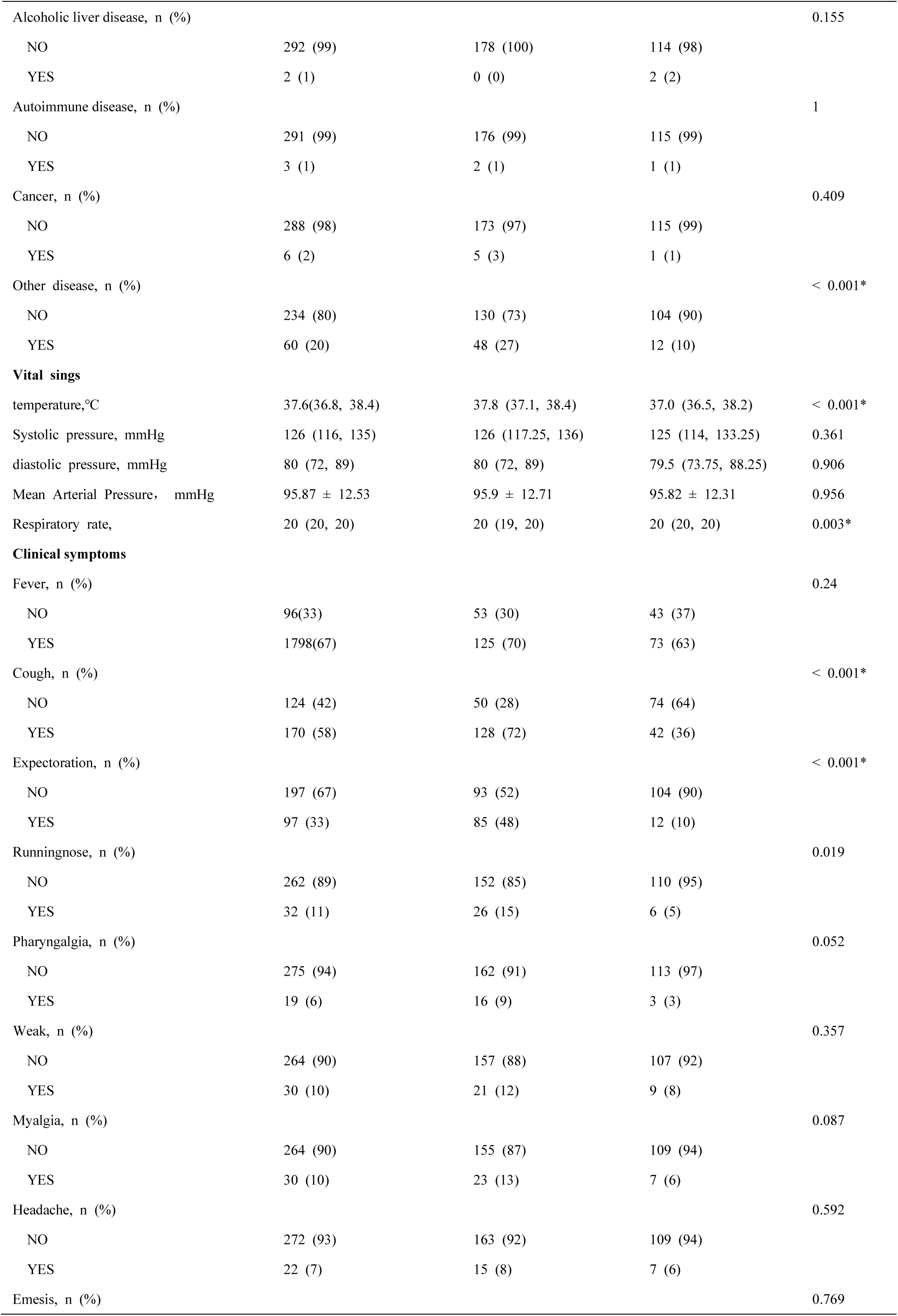
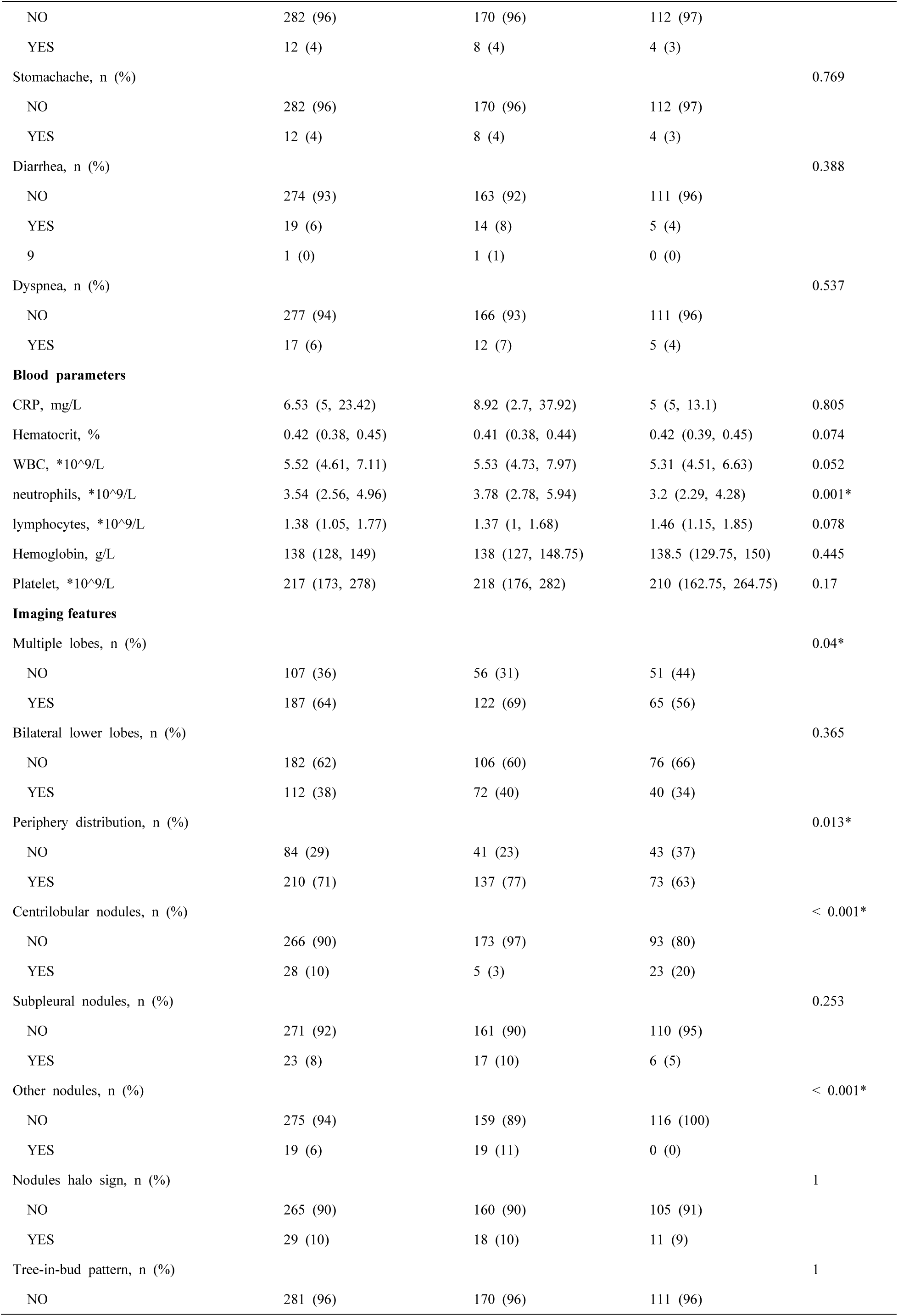
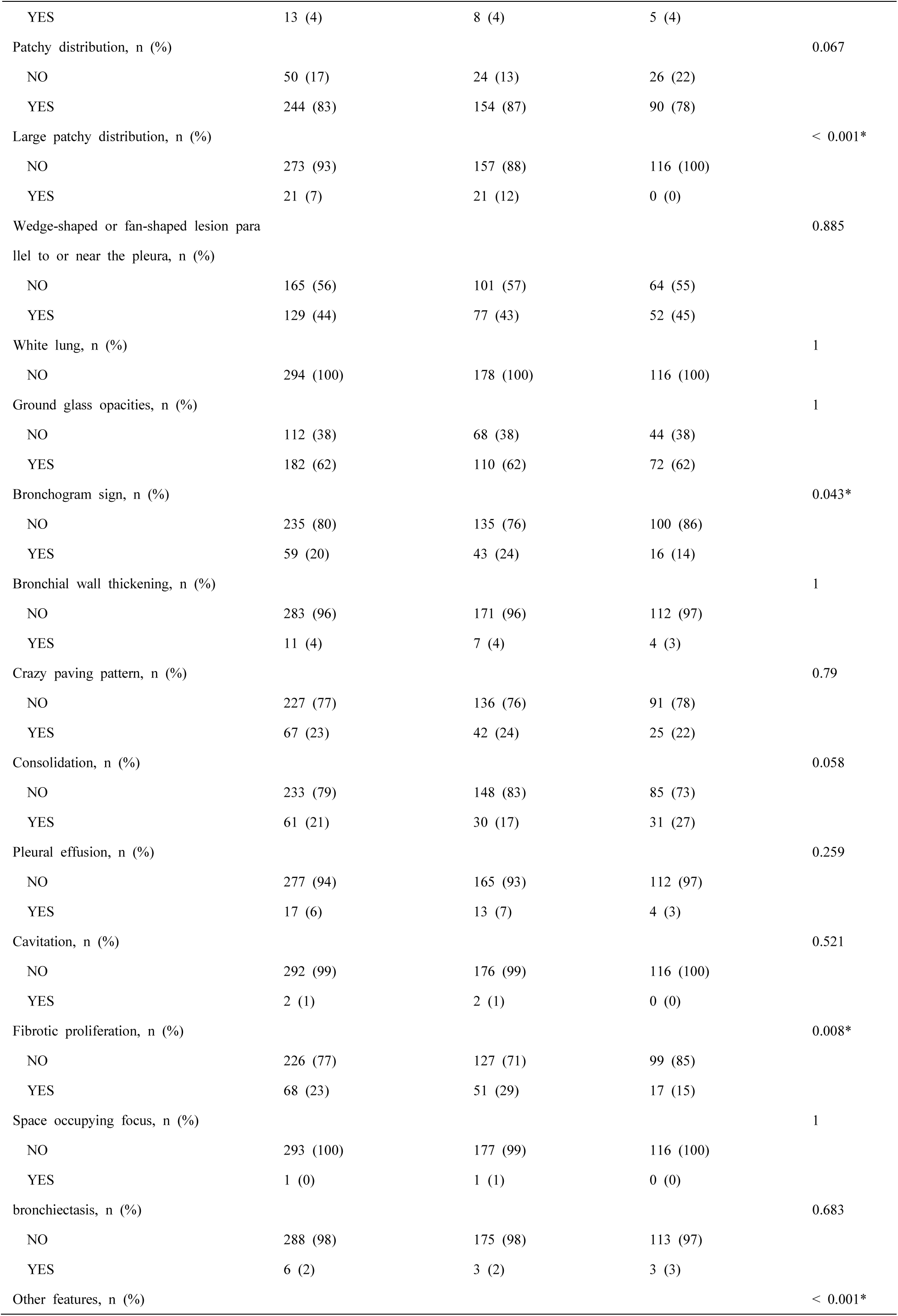
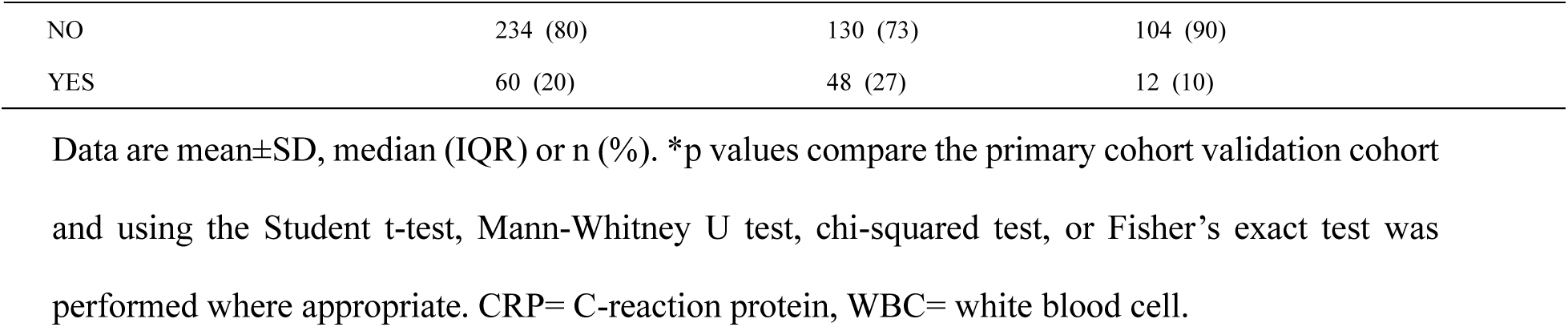
patients demographic, comorbidity, clinical symptoms, blood parameters and imaging features

| Variables | Total (n = 294) | primary cohort<br>(n = 178) | validation cohort<br>(n = 116) | *p* |
| --- | --- | --- | --- | --- |
| Age,years | 48 (35, 57) | 49.5 (37, 57.75) | 46 (35, 55.25) | 0.088 |
| Sex, n (%) |  |  |  | 0.87 |
| Male | 150 (51) | 92 (52) | 58 (50) |  |
| femal | 144 (49) | 86 (48) | 58 (50) |  |
| Drinking history, n (%) |  |  |  | 0.352 |
| NO | 283 (96) | 173 (97) | 110 (95) |  |
| YES | 11 (4) | 5 (3) | 6 (5) |  |
| Smoking history, n (%) |  |  |  | 0.029 |
| NO | 272 (93) | 170 (96) | 102 (88) |  |
| YES | 22 (7) | 8 (4) | 14 (12) |  |
| **Comorbidity** |  |  |  |  |
| Hypertension, n (%) |  |  |  | < 0.001* |
| NO | 243 (83) | 136 (76) | 107 (92) |  |
| YES | 51 (17) | 42 (24) | 9 (8) |  |
| diabetes, n (%) |  |  |  | 0.844 |
| NO | 261 (89) | 157 (88) | 104 (90) |  |
| YES | 33 (11) | 21 (12) | 12 (10) |  |
| COPD, n (%) |  |  |  | 0.564 |
| NO | 291 (99) | 177 (99) | 114 (98) |  |
| YES | 3 (1) | 1 (1) | 2 (2) |  |
| Chronic bronchitis, n (%) |  |  |  | 0.652 |
| NO | 289 (98) | 174 (98) | 115 (99) |  |
| YES | 5 (2) | 4 (2) | 1 (1) |  |
| bronchiectasis, n (%) |  |  |  | 0.395 |
| NO | 293 (100) | 178 (100) | 115 (99) |  |
| YES | 1 (0) | 0 (0) | 1 (1) |  |
| Asthma, n (%) |  |  |  | 0.281 |
| NO | 291 (99) | 175 (98) | 116 (100) |  |
| YES | 3 (1) | 3 (2) | 0 (0) |  |
| Tuberculosis, n (%) |  |  |  | 0.085 |
| NO | 288 (98) | 172 (97) | 116 (100) |  |
| YES | 6 (2) | 6 (3) | 0 (0) |  |
| Heart disease, n (%) |  |  |  | 0.669 |
| NO | 278 (95) | 167 (94) | 111 (96) |  |
| YES | 16 (5) | 11 (6) | 5 (4) |  |
| Heart failure , n (%) |  |  |  | 1 |
| NO | 286 (97) | 173 (97) | 113 (97) |  |
| YES | 8 (3) | 5 (3) | 3 (3) |  |
| Chronic nephritis, n (%) |  |  |  | 1 |
| NO | 292 (99) | 177 (99) | 115 (99) |  |
| YES | 2 (1) | 1 (1) | 1 (1) |  |
| Renal failure, n (%) |  |  |  | 0.156 |
| NO | 290 (99) | 174 (98) | 116 (100) |  |
| YES | 4 (1) | 4 (2) | 0 (0) |  |
| Liver cirrhosis, n (%) |  |  |  | 1 |
| NO | 291 (99) | 176 (99) | 115 (99) |  |
| YES | 3 (1) | 2 (1) | 1 (1) |  |
| Hepatic dysplasia, n (%) |  |  |  | 0.081 |
| NO | 289 (98) | 177 (99) | 112 (97) |  |
| YES | 5 (2) | 1 (1) | 4 (3) |  |
| Chronic hepatitis B, n (%) |  |  |  | 1 |
| NO | 281 (96) | 170 (96) | 111 (96) |  |
| YES | 13 (4) | 8 (4) | 5 (4) |  |
| Fatty liver, n (%) |  |  |  | 0.386 |
| NO | 289 (98) | 176 (99) | 113 (97) |  |
| YES | 5 (2) | 2 (1) | 3 (3) |  |
| Alcoholic liver disease, n (%) |  |  |  | 0.155 |
| NO | 292 (99) | 178 (100) | 114 (98) |  |
| YES | 2 (1) | 0 (0) | 2 (2) |  |
| Autoimmune disease, n (%) |  |  |  | 1 |
| NO | 291 (99) | 176 (99) | 115 (99) |  |
| YES | 3 (1) | 2 (1) | 1 (1) |  |
| Cancer, n (%) |  |  |  | 0.409 |
| NO | 288 (98) | 173 (97) | 115 (99) |  |
| YES | 6 (2) | 5 (3) | 1 (1) |  |
| Other disease, n (%) |  |  |  | < 0.001* |
| NO | 234 (80) | 130 (73) | 104 (90) |  |
| YES | 60 (20) | 48 (27) | 12 (10) |  |
| **Vital sings** |  |  |  |  |
| temperature,°C | 37.6(36.8, 38.4) | 37.8 (37.1, 38.4) | 37.0 (36.5, 38.2) | < 0.001* |
| Systolic pressure, mmHg | 126 (116, 135) | 126 (117.25, 136) | 125 (114, 133.25) | 0.361 |
| diastolic pressure, mmHg | 80 (72, 89) | 80 (72, 89) | 79.5 (73.75, 88.25) | 0.906 |
| Mean Arterial Pressure, mmHg | 95.87 ± 12.53 | 95.9 ± 12.71 | 95.82 ± 12.31 | 0.956 |
| Respiratory rate, | 20 (20, 20) | 20 (19, 20) | 20 (20, 20) | 0.003* |
| **Clinical symptoms** |  |  |  |  |
| Fever, n (%) |  |  |  | 0.24 |
| NO | 96(33) | 53 (30) | 43 (37) |  |
| YES | 1798(67) | 125 (70) | 73 (63) |  |
| Cough, n (%) |  |  |  | < 0.001* |
| NO | 124 (42) | 50 (28) | 74 (64) |  |
| YES | 170 (58) | 128 (72) | 42 (36) |  |
| Expectoration, n (%) |  |  |  | < 0.001* |
| NO | 197 (67) | 93 (52) | 104 (90) |  |
| YES | 97 (33) | 85 (48) | 12 (10) |  |
| Runningnose, n (%) |  |  |  | 0.019 |
| NO | 262 (89) | 152 (85) | 110 (95) |  |
| YES | 32 (11) | 26 (15) | 6 (5) |  |
| Pharyngalgia, n (%) |  |  |  | 0.052 |
| NO | 275 (94) | 162 (91) | 113 (97) |  |
| YES | 19 (6) | 16 (9) | 3 (3) |  |
| Weak, n (%) |  |  |  | 0.357 |
| NO | 264 (90) | 157 (88) | 107 (92) |  |
| YES | 30 (10) | 21 (12) | 9 (8) |  |
| Myalgia, n (%) |  |  |  | 0.087 |
| NO | 264 (90) | 155 (87) | 109 (94) |  |
| YES | 30 (10) | 23 (13) | 7 (6) |  |
| Headache, n (%) |  |  |  | 0.592 |
| NO | 272 (93) | 163 (92) | 109 (94) |  |
| YES | 22 (7) | 15 (8) | 7 (6) |  |
| Emesis, n (%) |  |  |  | 0.769 |
| NO | 282 (96) | 170 (96) | 112 (97) |  |
| YES | 12 (4) | 8 (4) | 4 (3) |  |
| Stomachache, n (%) |  |  |  | 0.769 |
| NO | 282 (96) | 170 (96) | 112 (97) |  |
| YES | 12 (4) | 8 (4) | 4 (3) |  |
| Diarrhea, n (%) |  |  |  | 0.388 |
| NO | 274 (93) | 163 (92) | 111 (96) |  |
| YES | 19 (6) | 14 (8) | 5 (4) |  |
| 9 | 1 (0) | 1 (1) | 0 (0) |  |
| Dyspnea, n (%) |  |  |  | 0.537 |
| NO | 277 (94) | 166 (93) | 111 (96) |  |
| YES | 17 (6) | 12 (7) | 5 (4) |  |
| **Blood parameters** |  |  |  |  |
| CRP, mg/L | 6.53 (5, 23.42) | 8.92 (2.7, 37.92) | 5 (5, 13.1) | 0.805 |
| Hematocrit, % | 0.42 (0.38, 0.45) | 0.41 (0.38, 0.44) | 0.42 (0.39, 0.45) | 0.074 |
| WBC, *10<sup>9</sup>/L | 5.52 (4.61, 7.11) | 5.53 (4.73, 7.97) | 5.31 (4.51, 6.63) | 0.052 |
| neutrophils, *10<sup>9</sup>/L | 3.54 (2.56, 4.96) | 3.78 (2.78, 5.94) | 3.2 (2.29, 4.28) | 0.001* |
| lymphocytes, *10<sup>9</sup>/L | 1.38 (1.05, 1.77) | 1.37 (1, 1.68) | 1.46 (1.15, 1.85) | 0.078 |
| Hemoglobin, g/L | 138 (128, 149) | 138 (127, 148.75) | 138.5 (129.75, 150) | 0.445 |
| Platelet, *10<sup>9</sup>/L | 217 (173, 278) | 218 (176, 282) | 210 (162.75, 264.75) | 0.17 |
| **Imaging features** |  |  |  |  |
| Multiple lobes, n (%) |  |  |  | 0.04* |
| NO | 107 (36) | 56 (31) | 51 (44) |  |
| YES | 187 (64) | 122 (69) | 65 (56) |  |
| Bilateral lower lobes, n (%) |  |  |  | 0.365 |
| NO | 182 (62) | 106 (60) | 76 (66) |  |
| YES | 112 (38) | 72 (40) | 40 (34) |  |
| Periphery distribution, n (%) |  |  |  | 0.013* |
| NO | 84 (29) | 41 (23) | 43 (37) |  |
| YES | 210 (71) | 137 (77) | 73 (63) |  |
| Centrilobular nodules, n (%) |  |  |  | < 0.001* |
| NO | 266 (90) | 173 (97) | 93 (80) |  |
| YES | 28 (10) | 5 (3) | 23 (20) |  |
| Subpleural nodules, n (%) |  |  |  | 0.253 |
| NO | 271 (92) | 161 (90) | 110 (95) |  |
| YES | 23 (8) | 17 (10) | 6 (5) |  |
| Other nodules, n (%) |  |  |  | < 0.001* |
| NO | 275 (94) | 159 (89) | 116 (100) |  |
| YES | 19 (6) | 19 (11) | 0 (0) |  |
| Nodules halo sign, n (%) |  |  |  | 1 |
| NO | 265 (90) | 160 (90) | 105 (91) |  |
| YES | 29 (10) | 18 (10) | 11 (9) |  |
| Tree-in-bud pattern, n (%) |  |  |  | 1 |
| NO | 281 (96) | 170 (96) | 111 (96) |  |
| YES | 13 (4) | 8 (4) | 5 (4) |  |
| Patchy distribution, n (%) |  |  |  | 0.067 |
| NO | 50 (17) | 24 (13) | 26 (22) |  |
| YES | 244 (83) | 154 (87) | 90 (78) |  |
| Large patchy distribution, n (%) |  |  |  | < 0.001* |
| NO | 273 (93) | 157 (88) | 116 (100) |  |
| YES | 21 (7) | 21 (12) | 0 (0) |  |
| Wedge-shaped or fan-shaped lesion parallel to or near the pleura, n (%) |  |  |  | 0.885 |
| NO | 165 (56) | 101 (57) | 64 (55) |  |
| YES | 129 (44) | 77 (43) | 52 (45) |  |
| White lung, n (%) |  |  |  | 1 |
| NO | 294 (100) | 178 (100) | 116 (100) |  |
| Ground glass opacities, n (%) |  |  |  | 1 |
| NO | 112 (38) | 68 (38) | 44 (38) |  |
| YES | 182 (62) | 110 (62) | 72 (62) |  |
| Bronchogram sign, n (%) |  |  |  | 0.043* |
| NO | 235 (80) | 135 (76) | 100 (86) |  |
| YES | 59 (20) | 43 (24) | 16 (14) |  |
| Bronchial wall thickening, n (%) |  |  |  | 1 |
| NO | 283 (96) | 171 (96) | 112 (97) |  |
| YES | 11 (4) | 7 (4) | 4 (3) |  |
| Crazy paving pattern, n (%) |  |  |  | 0.79 |
| NO | 227 (77) | 136 (76) | 91 (78) |  |
| YES | 67 (23) | 42 (24) | 25 (22) |  |
| Consolidation, n (%) |  |  |  | 0.058 |
| NO | 233 (79) | 148 (83) | 85 (73) |  |
| YES | 61 (21) | 30 (17) | 31 (27) |  |
| Pleural effusion, n (%) |  |  |  | 0.259 |
| NO | 277 (94) | 165 (93) | 112 (97) |  |
| YES | 17 (6) | 13 (7) | 4 (3) |  |
| Cavitation, n (%) |  |  |  | 0.521 |
| NO | 292 (99) | 176 (99) | 116 (100) |  |
| YES | 2 (1) | 2 (1) | 0 (0) |  |
| Fibrotic proliferation, n (%) |  |  |  | 0.008* |
| NO | 226 (77) | 127 (71) | 99 (85) |  |
| YES | 68 (23) | 51 (29) | 17 (15) |  |
| Space occupying focus, n (%) |  |  |  | 1 |
| NO | 293 (100) | 177 (99) | 116 (100) |  |
| YES | 1 (0) | 1 (1) | 0 (0) |  |
| bronchiectasis, n (%) |  |  |  | 0.683 |
| NO | 288 (98) | 175 (98) | 113 (97) |  |
| YES | 6 (2) | 3 (2) | 3 (3) |  |
| Other features, n (%) |  |  |  | < 0.001* |

|  |  |  |  |
| --- | --- | --- | --- |
| NO | 234 (80) | 130 (73) | 104 (90) |
| YES | 60 (20) | 48 (27) | 12 (10) |

Data are mean ± SD, median (IQR) or n (%). \*p values compare the primary cohort validation cohort and using the Student t-test, Mann-Whitney U test, chi-squared test, or Fisher's exact test was performed where appropriate. CRP= C-reaction protein, WBC= white blood cell.

Data are mean±SD, median (IQR) or n (%). *p values compare the primary cohort validation cohort and using the Student t-test, Mann-Whitney U test, chi-squared test, or Fisher’s exact test was performed where appropriate. CRP= C-reaction protein, WBC= white blood cell.

According to the univariate analysis (Supplementary materials), 16 related variables (gender, epidemiological history, two laboratory indicators: WBC and neutrophils count, twelve imaging features: multiple lobes, bilateral lower lobes, periphery distribution, nodules halo sign, patchy opacities, wedge-shaped or fan-shaped lesion parallel to or near the pleura, crazy paving pattern, ground glass opacities, consolidation, pleural effusion, air bronchogram sign and fibrotic proliferation) were included in the LASSO regression analysis(Figure 2). The results showed that gender, epidemiological history, neutrophils count, bilateral lower lobes, periphery distribution, wedge-shaped or fan-shaped lesion parallel to or near the pleura, ground glass opacities obtained from the primary cohort were predictive factors for COVID-19 pneumonia. According to the results of collinearity test and clinical practice, the epidemiological history, wedge-shaped or fan-shaped lesion parallel to or near the pleura, bilateral lower lobes, ground glass opacities, crazy paving pattern and WBC count were included in the final multivariate logistic regression analyses.

**Figure 2:**
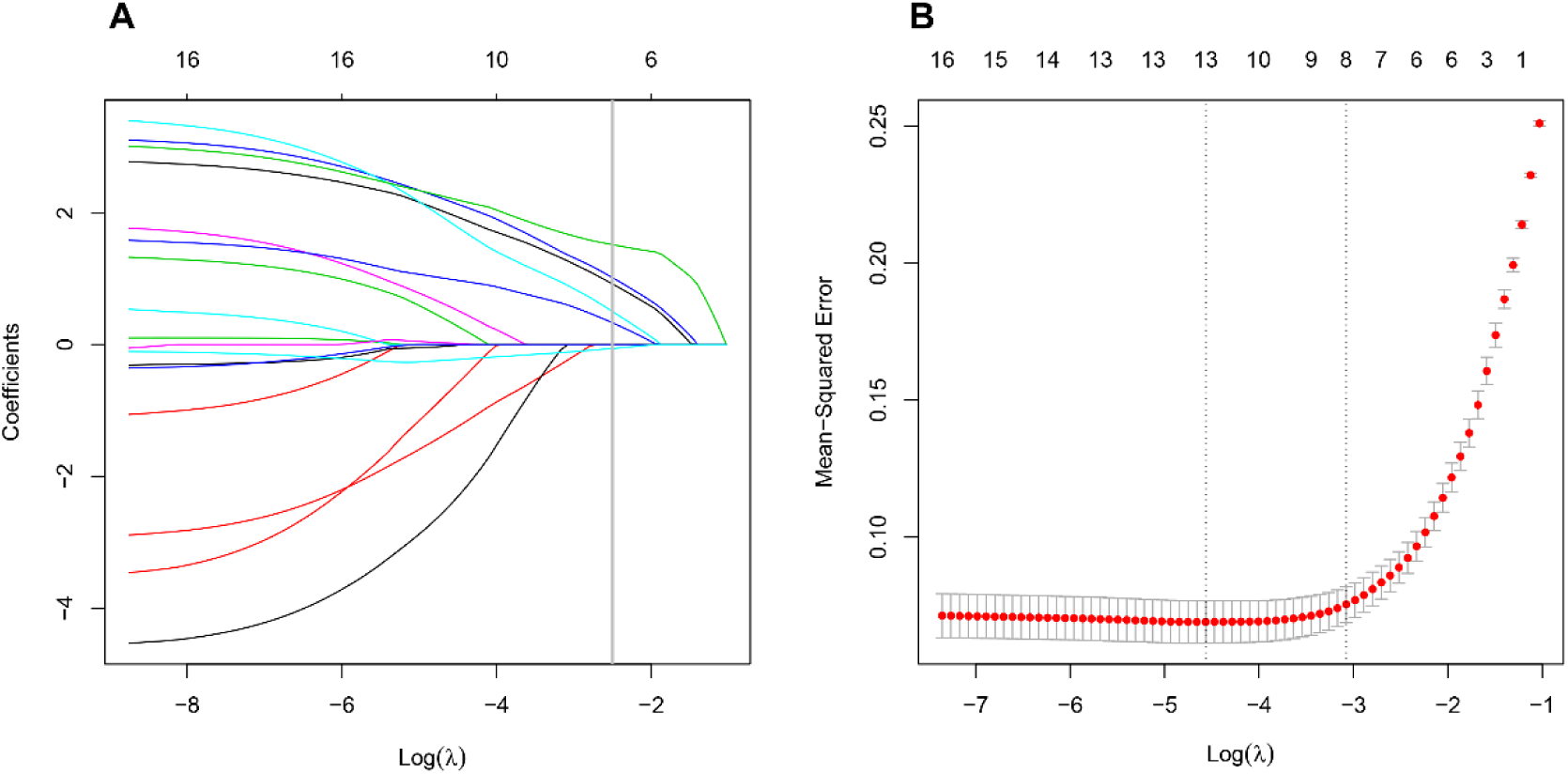
Predictive factors for COVID-19 pneumonia were selected by LASSO binary regression analyses. (A) The curve of the coefficient path of 16 predictive factors in primary cohort. The vertical line was set at the nonzero coefficients selected via 10-fold cross-validation, and there are 7 variables are included. (B) The adjustment penalty parameter *λ*was selected in the LASSO model by 10-fold cross-validation.

To provide a quantitative method to estimated COVID-19 pneumonia patients probability for clinician, we developed a nomogram (the ‘rms’ package was used in the R software). This nomogram had a C-statistic of 0.967 (95% confidence interval [CI], 0.943–0.992) for predicting the probability in suspected COVID-19 pneumonia patients (Figure 3). The radiomics nomogram was further internal validated using bootstrapping and external validated based on validation cohort. Good calibration curves were observed for the probability in suspected COVID-19 pneumonia patients in the internal validation data set and external validation cohort. The C-statistic of the radiomics nomogram were 0.848 in external validation cohort (Figure 4). Meanwhile, we developed another nomogram, based on initial viral nucleic acid detection, on the basis of univariable logistic analysis. The viral nucleic acid nomogram had a C-statistic of 0.961 (95% CI: 0.933–0.989) (Supplementary materials). Our nomogram showed higher discrimination, as compared with initial viral nucleic acid detection.

**Figure 3:**
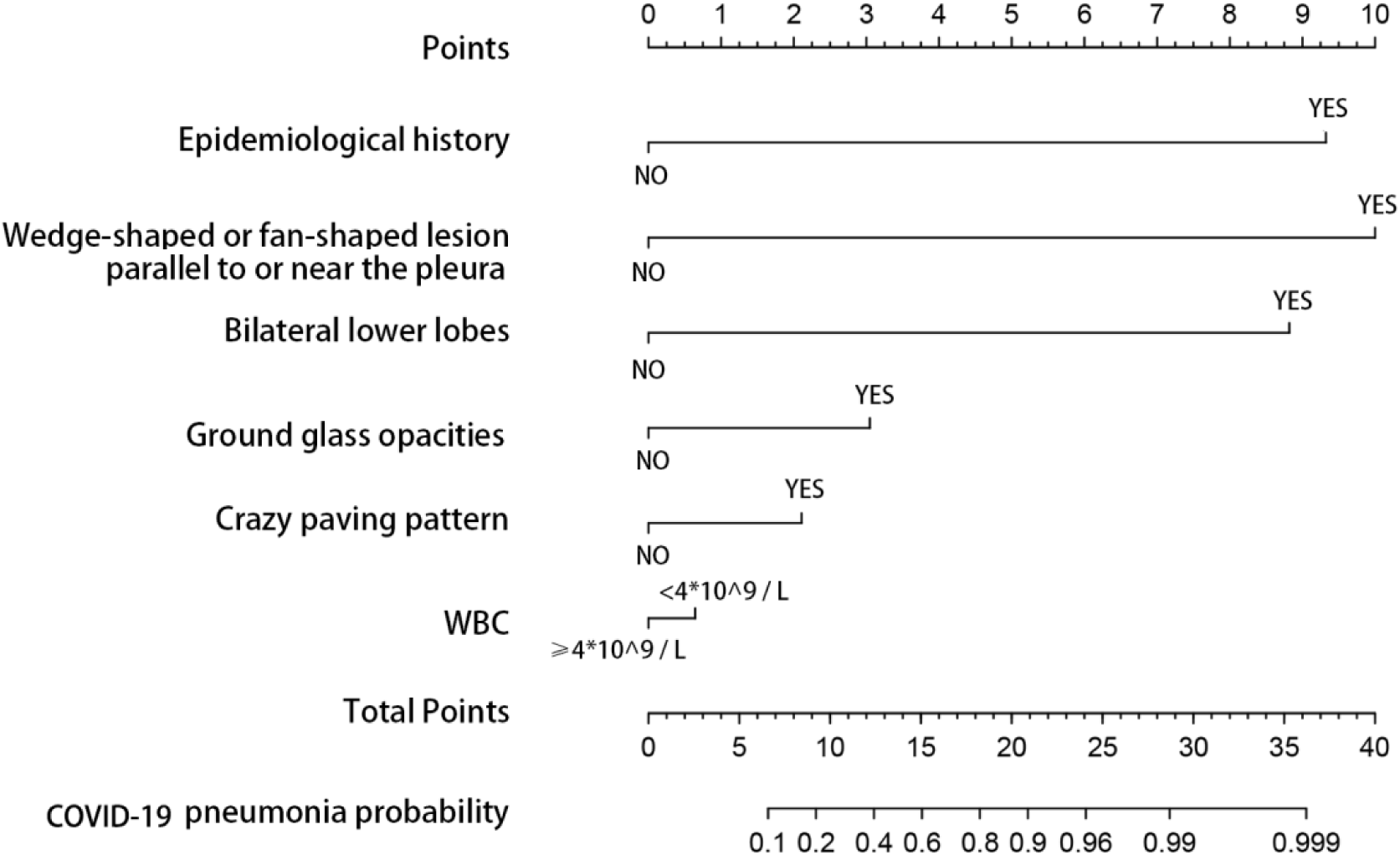
Developed radiomics nomogram. This nomogram was developed with four imaging features, epidemiological history and WBC count.

**Figure 4:**
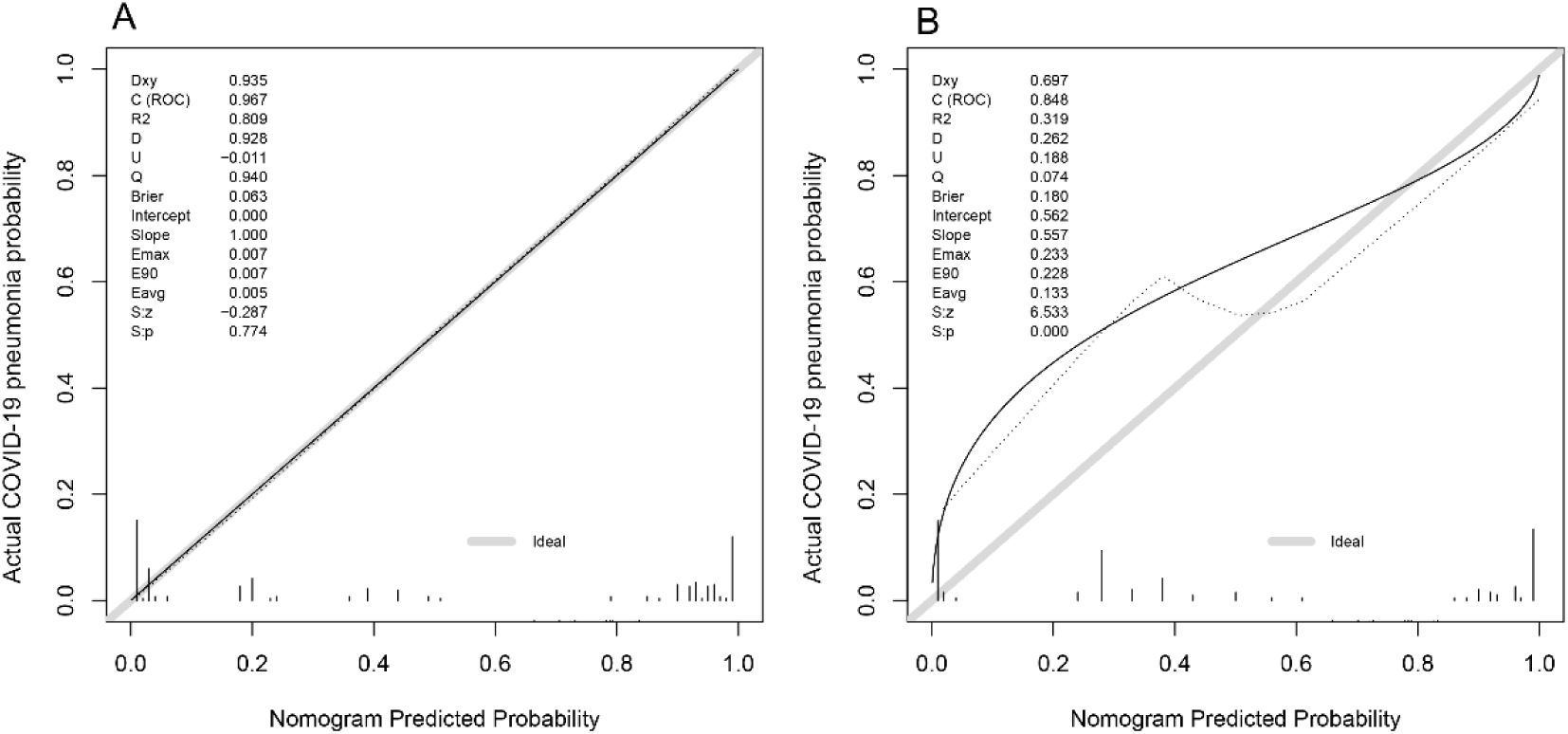
Calibration curves for predicting COVID-19 pneumonia probability by the nomogram in the in primary cohort (A) and validation cohort (B). Y-axis represents the actual COVID-19 pneumonia probability value, X-axis represents the predicted COVID-19 pneumonia probability value. Black solid line represents the prediction performance of the nomogram, the diagonal gray line represents an ideal nomogram model.

The DCA showed that radiomics nomogram had a very large threshold probability range, which means this nomogram had excellent net benefit to the outcome of suspected COVID-19 pneumonia patients. Clinical impact curve of the nomogram showed the predicted probability of COVID-19 pneumonia was very close to the actual COVID-19 pneumonia (Figure 5).

**Figure 5:**
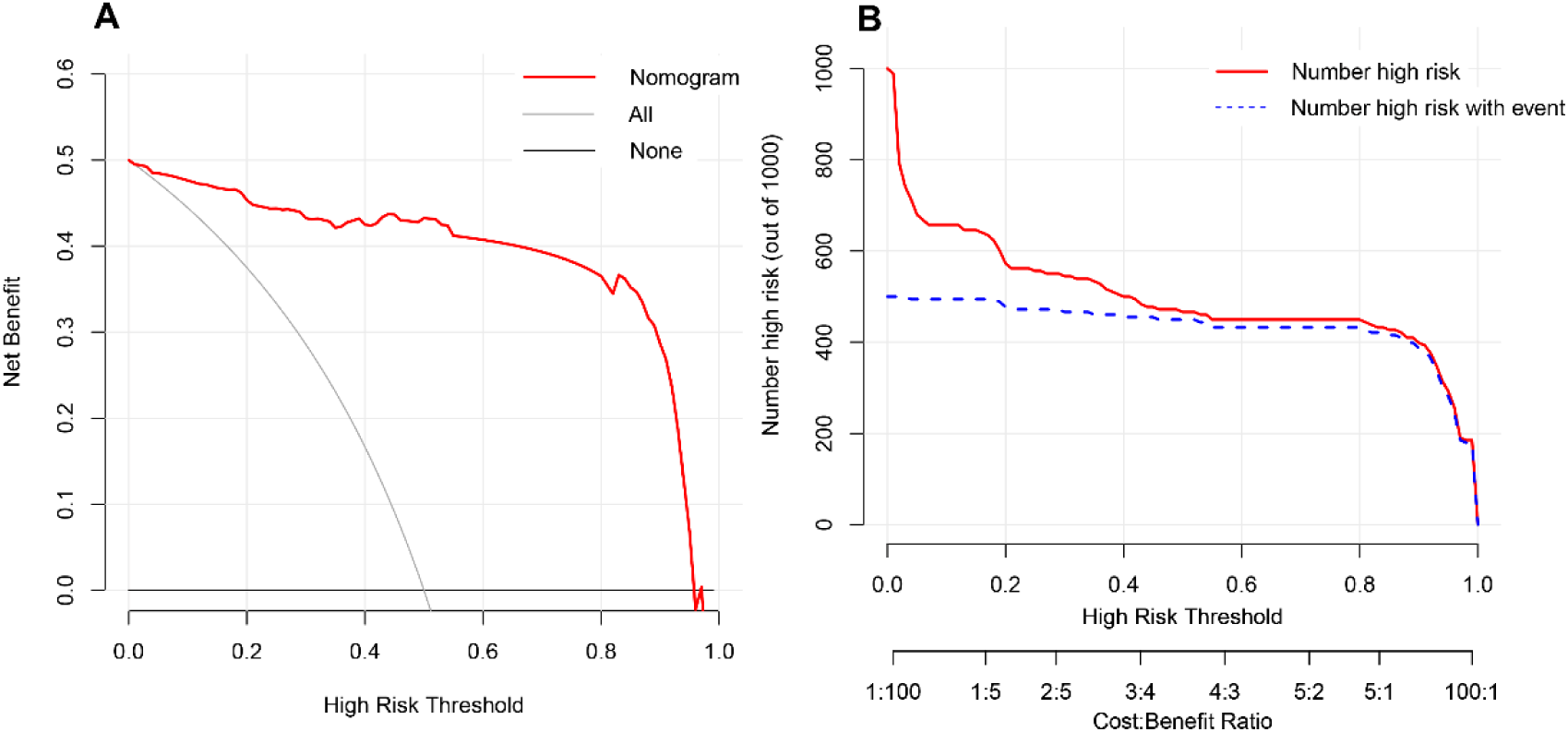
DCA of the nomogram model (A). The Y-axis represents the net benefit. The red line represents the predicted COVID-19 pneumonia nomogram model. Clinical impact curve of the nomogram model (B). The nomogram model is used to predict risk stratification for 1000 people. The red line represents the number of people classified high-risk by nomogram model under different threshold probability; the blue curve (Number high risk with event) is the number of truly positive people under different threshold probability.

## Discussion

In this study, we developed and validated a novel nomogram to predict COVID- 19 infection among patients who were suspected viral pneumonia. This diagnostic nomogram mainly relies on CT findings. Our study found that COVID-19 pneumonia is not significantly different from other suspected viral pneumonia in clinical symptoms and signs. There was also no significant difference in blood routine test, liver and kidney function test. Thus, according to clinical symptoms and signs and laboratory examinations, the COVID-19 pneumonia is difficult to distinguish from other viral pneumonia. However, they have similar and different manifestations on lung imaging detected by CT scan. ^10,17^ To this end, we have established this nomogram mainly based on lung imaging. All CT findings were analyzed from three aspects including distribution characteristics, morphology and density of pulmonary inflammation lesions.

The imaging features of viral pneumonia usually appear as multifocal ground glass opacities which correspond to pathological diffuse alveolar damage.^18^According to univariate analysis, ground glass opacities, crazy paving pattern, a wedge-shaped or fan-shaped lesion parallel to or near the pleura, the distribution characteristics of bilateral lower lobes and peripheral distribution of lesions are the characteristic imaging manifestation of COVID-19 pneumonia. But, the crazy paving pattern on the basis of multivariable was unassociated with COVID-19 pneumonia, which may be due to other potential confounding factors.^19^ But this does not mean that crazy paving pattern in COVID-19 pneumonia are unimportant. In addition, many studies have shown that the crazy paving pattern formed by interlobular septal thickening which was regarded as a typical imaging of viral pneumonia^13^. Therefore, we kept this factor in our model development. Another important imaging feature related to the characteristics of morphology and distribution, the wedge-shaped or fan-shaped lesion parallel to or near the pleura, which actually includes peripheral distribution of lesions. Because of their strong collinearity, it will seriously affect the accuracy of our research results^20^. Therefore, the imaging features of peripheral distribution was removed.

No matter from univariate or multivariate analysis results, epidemiological history plays a central role in the clinical diagnosis of COVID-19 pneumonia. Lymphocyte count did not show enough predictive strength. It may be related to the fact that these patients in our study are mainly suspected virus infection, and their lymphocyte count are generally low, with little difference. In view of the WBC of patients with viral infection is usually not high^21,22^, although it was unassociated in multivariate analysis, we still kept it in the process of establishing the diagnostic nomogram.

Finally, the nomogram incorporates 4 items of the imaging features, epidemiological contact history and WBC count status. Nomogram is a visualization of regression analysis, which is more and more widely used in clinical disease diagnosis, prognosis evaluation and efficacy evaluation^23-26^. Our results show that the nomogram based on imaging features has good sensitivity and specificity in the diagnosis of COVID-19 pneumonia. Moreover, its discrimination for COVID-19 pneumonia is better than the first detection of viral nucleic acid. If there is a lack of virus nucleic acid test kit, COVID-19 pneumonia can be determined by lung CT preferentially.

In order to prove the calibration of the nomogram, clinical data was collected from different institutions. As is well known, the internal validity associated with the explanation of the results, and the external validity related to the generalizability of the results^27,28^. Through the internal and external validation data set analysis, the calibration of our nomogram has been proved to be highly consistent. This means that our nomogram may be popularized and applied widely in other hospital. However, to evaluate its clinical usefulness, it depends on how much it benefits the patient, not just its popularization^29^. DCA is an novel method^30,31^, it offers insight into clinical consequences on the basis of threshold probability, from which the net benefit could be derived^32^. The DCA showed that if we choose to diagnose COVID-19 pneumonia with a 60% threshold probability, 40 out of every 100 people will benefit.

### Limitations of this study

Our study has several limitations. Firstly, only 178 patients were included in primary cohort and another hospital was selected for external validation (116 patients). Whether this nomogram is applicable to patients with other areas background is still unclear. A large number of patients as data need to be collected to verify its clinical application. Secondly, this nomogram is mainly used to identify COVID-19 pneumonia in the patients with suspected viral pneumonia, not all types of pneumonia. Although the decrease of lymphocyte count is more common among COVID-19 pneumonia, not observed in our study. It may be related to our inclusion criteria.

## Conclusion

In conclusion, this study presents a novel nomogram that incorporates both the imaging features, epidemiological history and WBC. It can predict COVID-19 pneumonia conveniently and accurately. Using this nomogram has high net benefit for patients with suspected COVID-19 infection.

## Data Availability

All data referred to in the manuscript is available.

## AUTHOR CONTRIBUTION

Conception and design: Zhiyi Wang, Daqing Chen, Liang Wang, Xiaodong Liu, Xian Shen, Shengwei Jin

Collection and assembly of data: Jie weng, Zhongwang Li, Ruonan Hou, Lebin Zhou,Hua Ye,Ying Chen, Ting Yang

Zhiyi Wang^1,2^*#, Jie Weng^1^*, Zhongwang Li^3^, Ruonan Hou^1^, Lebin Zhou^4^, Hua Ye^4^,

Ying Chen^1^, Ting Yang^1^, Daqing Chen^1^, Liang Wang^5^, Xiaodong Liu^2^, Xian Shen^6^#, Shengwei Jin^3^#

